# Feasibility and safety of long-term remote-only monitoring in a large cohort of pacemaker patients

**DOI:** 10.64898/2026.03.12.26348292

**Authors:** Markus Sane, Toni Jäntti, Annukka Marjamaa, Elina Pennanen, Charlotte Aura, Eeva Torvinen, Leena Karjalainen, Jarkko Karvonen, Pekka Raatikainen

## Abstract

**Background:** Remote monitoring (RM) enables convenient follow-up of patients with cardiac implantable electronic devices. In remote-only monitoring (RM-only) in-person visits are only scheduled if needed based on RM findings. This study evaluated the feasibility and safety of long-term RM-only of pacemaker (PM) patients.

**Methods:** All patients with Biotronik PM were included in the analysis. Data on the number and causes of additional in-office device interrogations, actions due to the transmissions, hospitalizations and performance of RM were collected from a large cohort of pacemaker patients followed by RM-only.

**Results:** In total 606 patients (302 female) with mean age of 78±12 years were included in the analysis. During the mean follow-up of 2.8 years 445 additional in-office device interrogations were made in 287 patients (0.3 interrogations / year), and in 110 (25%) of these cases changes to device programming were made. In a subgroup analysis of 100 patients with at least one year of prior appointment-based device monitoring, the need for in-office visits was 6.9 times higher per follow-up year than in RM (IRR=6.9, 95% CI 4.9-9.9; p<0.001). The hospitalization rate in the entire cohort during RM was 0.3 / year with no difference in the rate of hospitalizations between the two monitoring methods (IRR 1, 95% CI 0.8-1.4, p=0.8). The success of daily transmissions was 91.7 %.

**Conclusion:** Our real-word data indicate that RM-only offers an efficient and safe method for long-term follow-up of pacemaker patients.

## Introduction

The ability to remotely follow cardiac implantable electronic devices (CIED) has improved patient care and reduced the workload of the device clinics without compromising safety (^1–3^). Remote monitoring (RM) is associated with reduced need for in-person device interrogations, lower hospital staff workload, near real-time detection of device-related adverse events and tachyarrhythmias such as atrial fibrillation or ventricular tachycardia and improved patient adherence to the therapy ^(1–7)^. Thus, RM reduces health care costs and increases the quality of life of the patients ^(8)^.

According to the current guidelines during RM the intervals between additional in-office visits can be prolonged but the visits cannot be completely omitted ^(9)^. Remote-only (RM-only) device follow-up was increasingly used during the COVID pandemic, and the European Society of Cardiology recommended that routine device interrogations in the hospitals should be replaced by RM whenever possible (^10^). Recently, two studies have compared RM-based follow-up without scheduled in-patient visits to conventional in-office CIED follow-up. In the At-home study 1274 patients were randomized to conventional (twice yearly in-office visits) and remote-only follow-up. During a two-year follow-up there was no difference in the primary composite end-point of death, stroke, or cardiovascular events requiring surgery (^11^). In keeping with this, the results of the RM-ALONE study in which 445 patients were randomized to either remote-only follow-up or remote follow-up with additional in-office visits every 6 months found no difference in major adverse cardiovascular events between the two groups during a two-year study period (^12^). In addition, it has been reported that patients have positive attitude towards using RM as the primary follow-up method (^13^). These findings suggest that follow-up by RM-only is feasible, but data on long-term follow-up in a real-world setting is lacking.

The aim of this study was to evaluate the impact of long-term follow-up by RM-only on the workload, patient safety and the performance of RM technology in a large cohort of pacemaker (PM) patients.

## Methods

This study was approved by the institutional review board of Helsinki University Hospital. Following consultation with the ethics committee, it was determined that ethics permission was not required.

### Patient population

The Helsinki University Hospital Heart and Lung Center is a tertiary care center with a total catchment population of 2.1 million inhabitants. In our clinic, RM has been the primary choice for high energy device follow-up for more than a decade and since 2020 the follow-up of pacemaker patients has been performed exclusively with RM after the initial in-office visit 1−3 months post-implantation. Currently, we take care of more than 4000 CIED patient remote follow-up (Supplementary Table S1).

At the end of 2024 a total of 1956 pacemaker patients had been enrolled to our RM system. However, only those using Biotronik RM (Home-Monitoring®, Biotronik SE and Co. KG, Berlin, Germany) were included in this study for the following reasons. First, in the past years Biotronik has been our main supplier of PMs and we initiated RM for Biotronik PMs in spring 2020 and therefore the longest follow-up is available for these devices. Second, unlike the other RM systems Home-Monitoring® transmits technical and medical data daily on a secure internet site enabling evaluation of daily success of RM transmissions. The date of the last device interrogation is also available in the portal. If the date of the last device interrogation in Home monitoring® portal was the same as the date of RM initiation it was determined that the patient has been followed solely with RM.

In total 1078 patients with Biotronik PMs were followed remotely in our clinic at the end of 2024 and were included in the RM workload analysis. The patients (n=606) in whom RM was initiated before December 31, 2022 were included in the analysis concerning the feasibility, safety and performance of RM-only approach. A subgroup of 100 patients with more than one year of appointment-based monitoring before RM initiation was used to compare the workload and safety between these two monitoring methods. The flowchart of the patients included in this study is presented in Figure 1.

**Figure 1.**
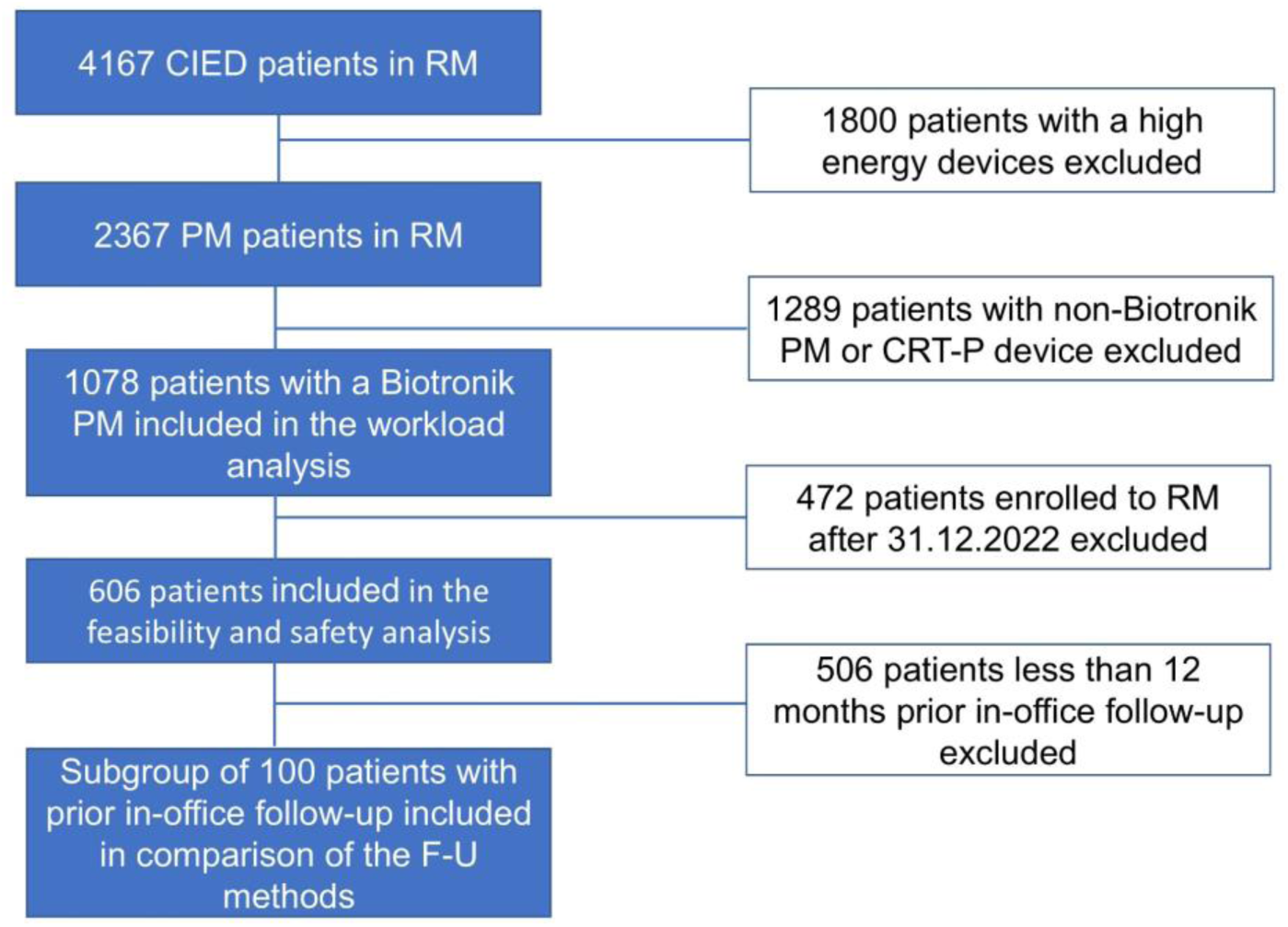
Flow diagram for inclusion and exclusion of the participants. Detailed information on the maufacturer and type of the devices is presented in the Supplementary Table S1.

All patients received oral and written information on the benefits and potential drawbacks of CIED follow-up options and RM was initiated if patients gave oral consent.

### Data collection

Our remote monitoring database has been presented earlier (^14^). Briefly the data on the manufacturer, type of device, serial number, reason, and actions after each transmission was collected. In addition, the data was collected from the Biotronik Home-Monitoring® database and the hospital electronic health records (EHR). The daily success rate of the RM transmissions was defined as the proportion of daily transmissions that were transmitted to the Home-Monitoring® portal across all days. The workload in RM per PM was calculated by dividing the number of transmissions with the mean number of Biotronik PMs in RM in 2023 and 2024. Noteworthy, the data on drop-out from the RM was available since 1^st^ of March 2024 and this data was collected from the total cohort of Biotronik PM patients in RM until the end of February 2025.

#### Device programming

The devices were programmed individually by the implanting electrophysiologist. In general, the nominal values were used for the automatic threshold, sensitivity and blanking periods and atrioventricular (AV) delay of 180 ms after paced and 140 ms after sensed atrial event were used with AV-hysteresis of +110 ms for patients with intact atrioventricular conduction. Activity sensor was programmed on if there were signs of sinus node dysfunction.

### RM settings

We used a previously published approach for the RM alert settings (^14^). This encompasses both the preprogramming of alert settings and the modification of alert settings after alert transmissions to reduce the number of unnecessary transmissions. Lead integrity alerts were turned on with normal impedance range of 200-1500 Ω and pacing threshold cut-off was > 3.0 V / 0.4 ms for both the atrial and the ventricular lead. Sensing alert trigger was programmed at <0.2 mV for the atrial and <2.0 mV for the ventricular lead, respectively. More detailed information on the settings of the RM alerts is presented in the Supplementary appendix Figure S1 A-C

In addition to the alert transmissions, annual scheduled transmissions were used in all patients. The scheduled transmissions were evaluated and documented in the EHR as suggested in the recent consensus statement ^(15)^, and the summary of the findings was also available for the patients via nationwide EHR.

Alert transmission for high ventricular rate (HVR) were nominally turned off in all patients and HVR episodes were assessed from the scheduled follow-up transmissions, with the trigger for HVR episodes programmed to the lowest count. Atrial fibrillation (AF) alerts were turned off in anticoagulated patients, and in non-anticoagulated patients we used 25 % AF burden as the alert trigger irrespective of the individual CHA_2_DS_2_-VA score. The trigger for high ventricular rate during AF was set to mean heart rate > 100 bpm per day. If persistent AF of more than six months of duration was detected in the scheduled transmission and no cardioversion was planned, an in-office visit was arranged to program an appropriate permanent pacing mode.

The threshold for RM disconnection was set to 21 days and patients were advised that if they were away from home less than 3 weeks, they can travel without RM transmitter and with longer durations out of home they were asked to inform the device clinic beforehand.

In our RM workflow nurses scan the RM portals every business day and analyze all the annual and patient-initiated transmissions and alert-transmissions are analyzed either by nurse or electrophysiologist. Annual transmissions are evaluated within two weeks and alert and patient-initiated transmissions on the same business day. Nurses also contact patients if disconnection episodes have occurred and ensure that connectivity is restored. The patient can freely contact device clinic during business hours. Nurses consult electrophysiologists when needed, and all consultations are conducted through the messaging tools integrated into the EHR system.

### Statistics

The data was analyzed with IBM SPSS 25.0 for Windows (IBM Corp., Armonk, NY, USA). The normality of continuous variables was assessed by the Kolmogorov–Smirnov test. Data are presented as means (with standard deviation for normally distributed continuous variables) and as median (with interquartile range when distribution was skewed). Mc Nemars test was used to evaluate the need of ER evaluations or hospitalizations in a subgroup of patients with both in-office follow-up and RM. Negative binomial regression test was used to evaluate the rate of device interrogations, ER evaluations or hospitalizations between the two follow-up methods in this subgroup.

The statistical significance level was set to 5%.

## Results

A total of 1078 patients with a Biotronik PM were included in the study. In 606 of these patients (302 female) RM was initiated before December 31, 2022 allowing an average of 3 years (1.7-4.6 years) of follow-up by RM-only.

The patient characteristics are presented in Table 1. The detailed information on the PM type is provided in Supplementary Table S2

**Table 1.**
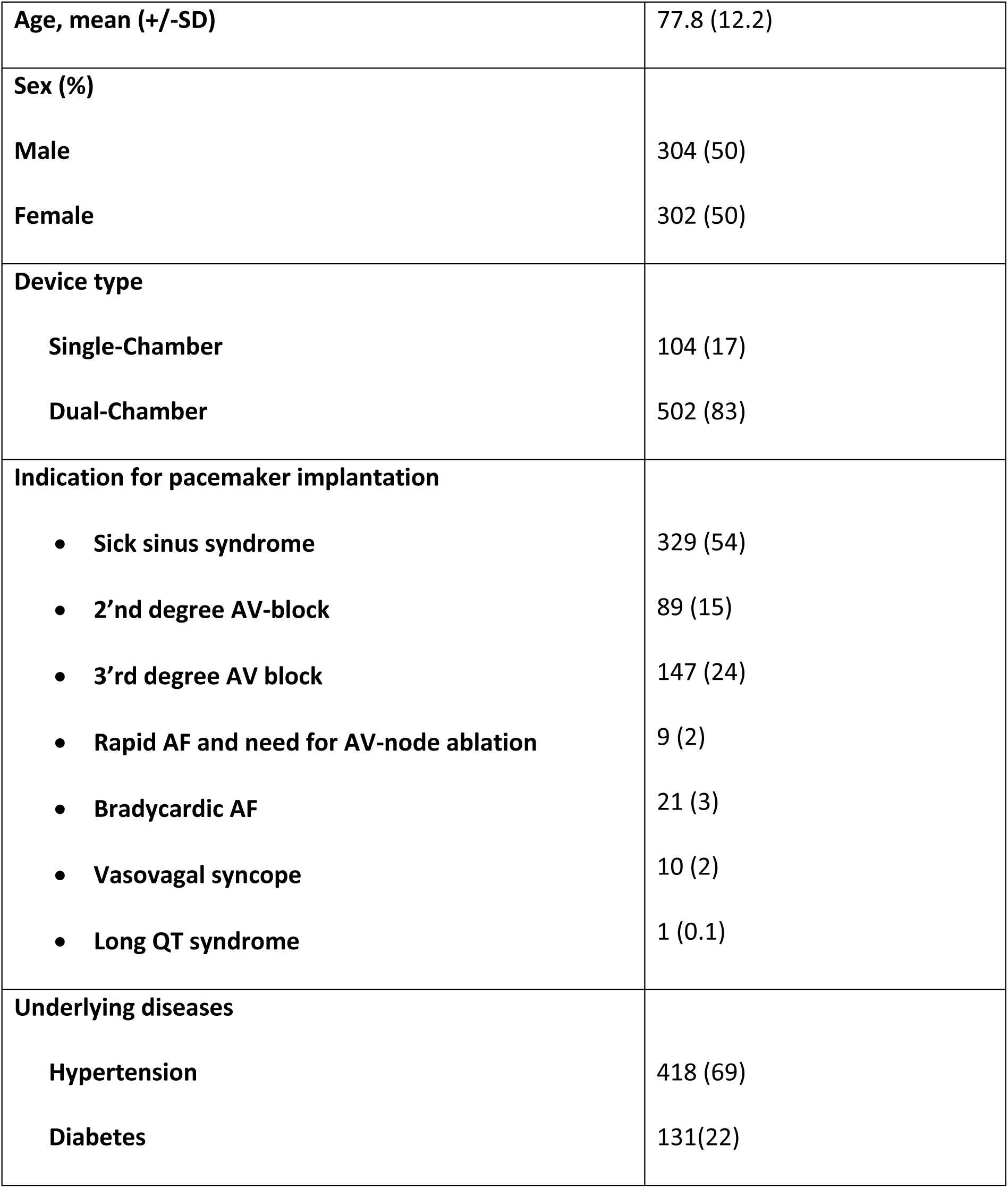
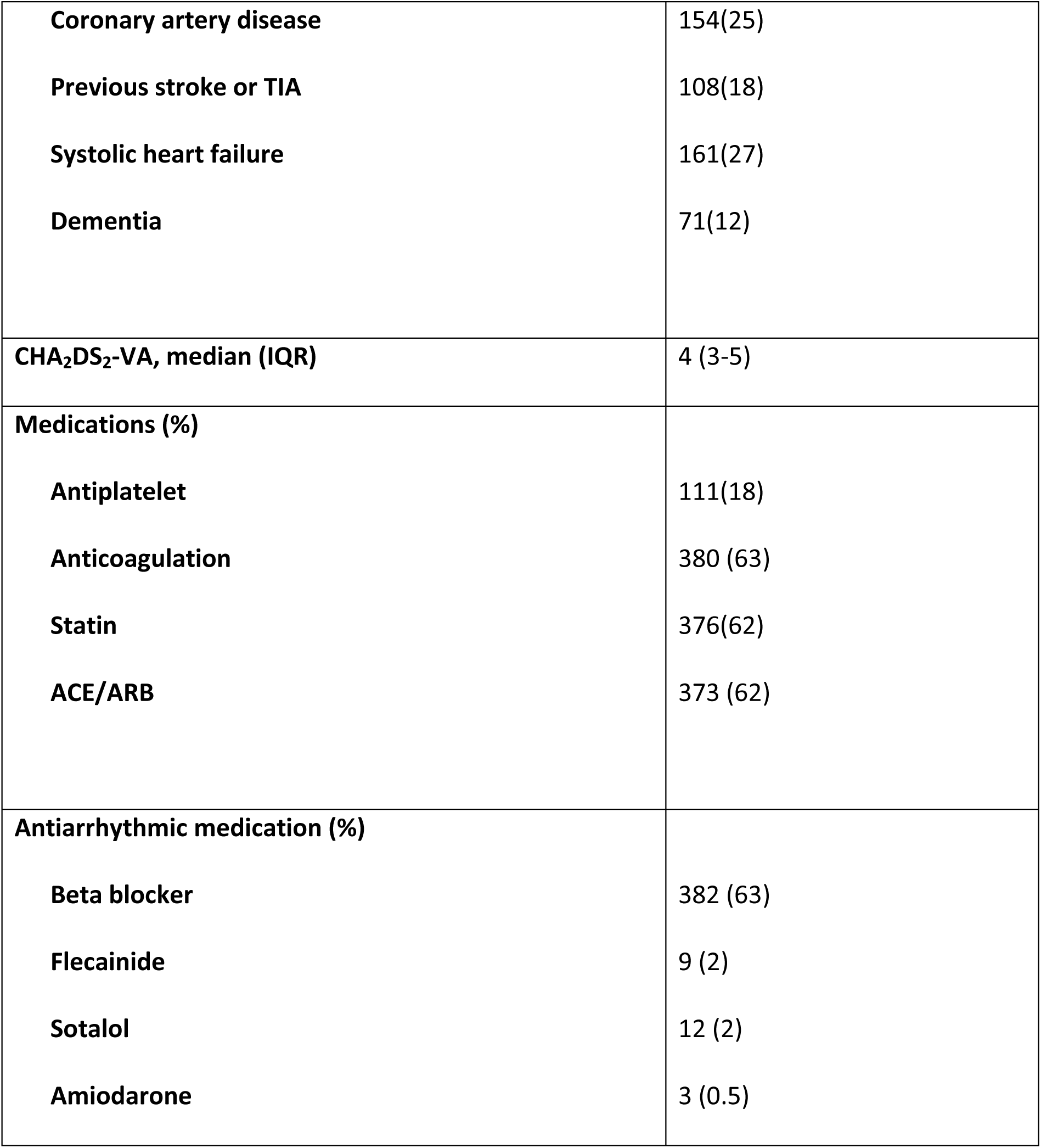
Patient characteristics.

|  |  |
| --- | --- |
| **Age, mean (+/-SD)** | 77.8 (12.2) |
| **Sex (%)** |  |
| Male | 304 (50) |
| Female | 302 (50) |
| **Device type** |  |
| Single-Chamber | 104 (17) |
| Dual-Chamber | 502 (83) |
| **Indication for pacemaker implantation** |  |
| • Sick sinus syndrome | 329 (54) |
| • 2<sup>nd</sup> degree AV-block | 89 (15) |
| • 3<sup>rd</sup> degree AV block | 147 (24) |
| • Rapid AF and need for AV-node ablation | 9 (2) |
| • Bradycardic AF | 21 (3) |
| • Vasovagal syncope | 10 (2) |
| • Long QT syndrome | 1 (0.1) |
| **Underlying diseases** |  |
| Hypertension | 418 (69) |
| Diabetes | 131(22) |
| **Coronary artery disease** | 154(25) |
| **Previous stroke or TIA** | 108(18) |
| **Systolic heart failure** | 161(27) |
| **Dementia** | 71(12) |
| **CHA<sub>2</sub>DS<sub>2</sub>-VA, median (IQR)** | 4 (3-5) |
| **Medications (%)** |  |
| **Antiplatelet** | 111(18) |
| **Anticoagulation** | 380 (63) |
| **Statin** | 376(62) |
| **ACE/ARB** | 373 (62) |
| **Antiarrhythmic medication (%)** |  |
| **Beta blocker** | 382 (63) |
| **Flecainide** | 9 (2) |
| **Sotalol** | 12 (2) |
| **Amiodarone** | 3 (0.5) |

**Table 2.** The reasons for in-office device interrogation during remote monitoring.

| In-office device interrogations during remote monitoring-based follow-up |  |  |  |  |  |
| --- | --- | --- | --- | --- | --- |
| Reason | First<br>interrogation | Second<br>interrogation | Third<br>interrogation | Fourth<br>interrogation | Total<br>number<br>(% of<br>all) |
| Due to finding in RM<br>transmission* | 71 | 22 | 12 | 7 | 112<br>(27) |
| Patient request | 61 | 17 | 12 | 9 | 99 (23) |
| Request from another<br>department or<br>cardiologist | 23 | 3 | 3 |  | 29 (7) |
| MRI | 48 | 16 | 2 | 2 | 68 (16) |
| Surgery or other | 18 | 7 | 4 |  | 29 (7) |

| **procedure** |  |  |  |  |  |
| --- | --- | --- | --- | --- | --- |
| **Radiation therapy** | 11 | 4 | 4 | 1 | 20 (5) |
| **Electrical cardioversion** | 4 | 2 |  |  | 6 (1) |
| **Lack of EGM in RM due to older PM type** | 12 | 1 |  |  | 13 (3) |
| **Disconnection of RM** | 17 | 2 | 2 |  | 21 (5) |
| **System error\*\*** | 19 | 3 |  |  | 22 (5) |
| **No data available\*\*\*** | 3 |  |  |  | 3 (1) |
| **Total number\*\*\*\*** | 287 | 77 | 40 | 20 | 422<br>(100) |

\*40 were due to permanent AF and mode switch and 45 were due to failed threshold test and subsequent scheduled threshold testing
\*\*Negligent error of forgetting to program the home monitoring On or programming the interval for scheduled transmissions to 30 days instead of 180 days in ENITRA 6 devices.
\*\*\* *Home-Monitoring*® shows that device has been interrogated but no reason can be found from the electronic health records.
\*\*\*\* Eleven patients had more than 4 device interrogations and in total these patients had 23 additional interrogations due to various causes

### In-office device interrogations during remote monitoring

The need for additional in-office device interrogation during RM occurred at a rate of 0.3 interrogations per follow-up year. In a subgroup of patients with a history of both monitoring methods the mean duration of appointment-based and RM-only follow-up was 2.8 and 3.0 years, respectively (p=0.3). The need for in-office visits was 6.9 times higher per follow-up year during appointment-based than in RM-only follow-up (IRR=6.9, 95% CI 4.9-9.9; p<0.001).

During follow-up 319 (53%) patients needed no in-office visits, whereas 287 (47%) patients had at least one in-office device interrogation identified from our EHR or Home Monitoring® portal. In total 442 device interrogations were documented in EHR but the data of three interrogations (0.7%) was lacking, and these were only identified from Home Monitoring® portal. The median number of in-office device interrogations in these patients was one (IQR 1-2).

### The need for programming changes

Any changes to device programming were made in 110 (18%) patients. The most common programming adjustments included change in the pacing mode due to permanent AF (n=40, 36%), adjustments to activity sensor settings (n=34, 31%), change in atrioventricular conduction delay (n=11, 10%), programming automatic threshold test to monitoring (n=4, 4%), change in lower or upper pacing rate (n=4, 4%). Multiple programming changes during interrogation were done in 17 patients (15%). The median time for programming changes after the initiation of RM was 530 days (IQR 222-845 days).

### Performance of RM

The daily success of RM transmissions was on average 91.7% (SD 10.1%) and in 47 patients (8%) the rate was below 75%.

RM was disconnected at least once in 185 (30.5%) patients and the median number of disconnections in these patients was one (IQR 1-2). In total 379 disconnections episodes were registered from Home-Monitoring® (0.2 events/year). Twenty-one patients (3%) needed in-office visit to restore the RM connection.

### Workload related to RM

Overall, 2553 RM transmissions (2061 scheduled- and 492 alert-transmissions) were generated during 2023 and 2024 corresponding to a rate of 1.3 / device/ year. Majority of these were non-actionable (2242, 88%). RM alerts occurred in 242 patients (40%). The triggers, actions taken, and rate of alert transmissions for 2023 and 2024 as well as the data on scheduled transmission is presented in the Supplementary Table S3A. The most comment triggers for the alert transmissions were high ventricular rate during AF and atrial high rate episode (AHRE).

### Safety

During the follow-up 342 patients were evaluated in emergency room (ER) at least once and the total number of ER visits was 946 (median 1, IQR 0-2). The median number of ER visits was 0.3 / year (IQR 0-0.8). Among these patients the ER evaluation led to hospitalizations at least once in 277 patients and the overall number of hospitalizations in these patients was 667 (median 2, IQR 1-3, rate 0.3 / year) and median duration of hospitalizations was 6 days (IQR 3-18). The most common causes for hospitalizations are presented in Table 3.

**Table 3.**
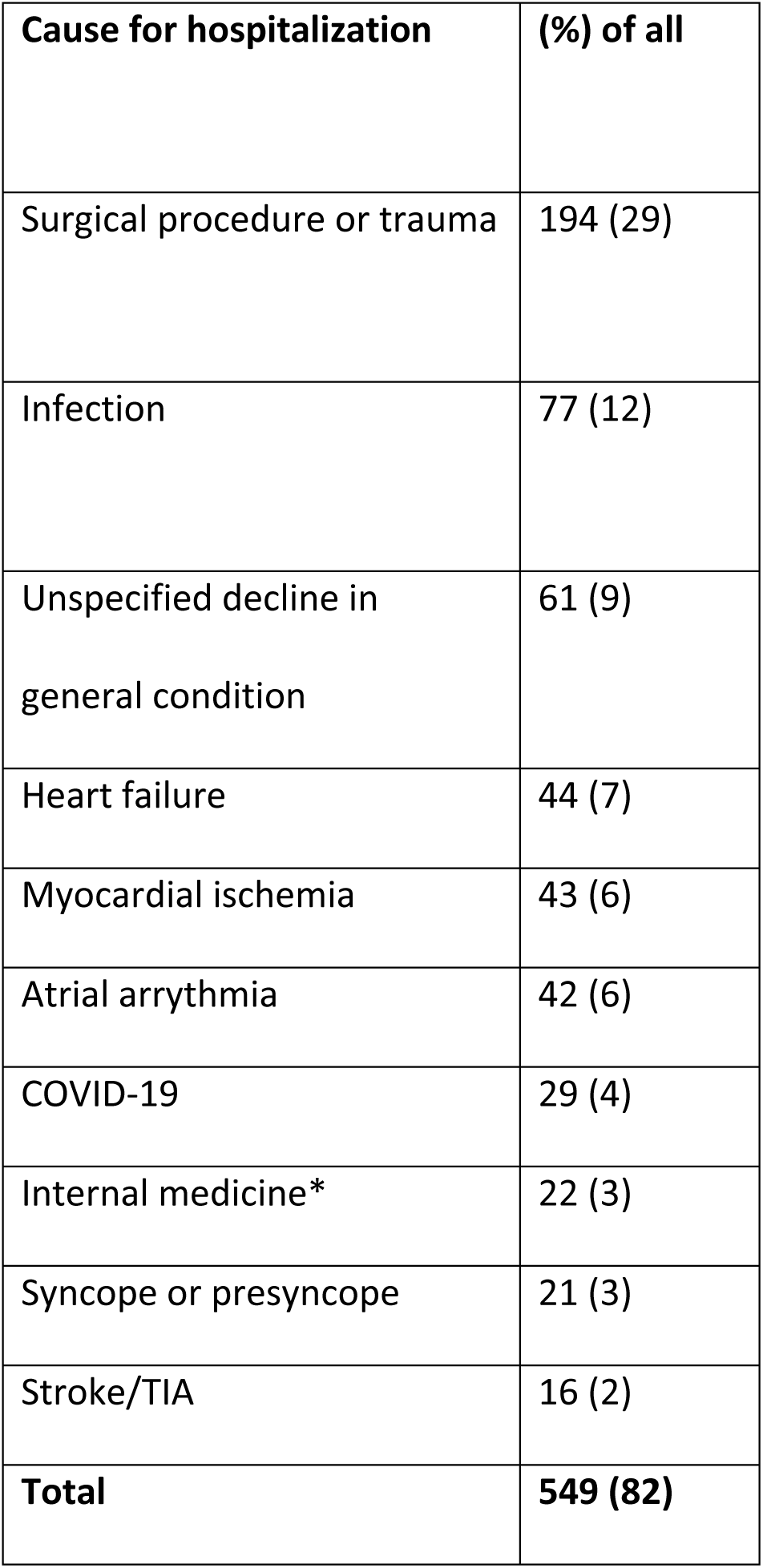
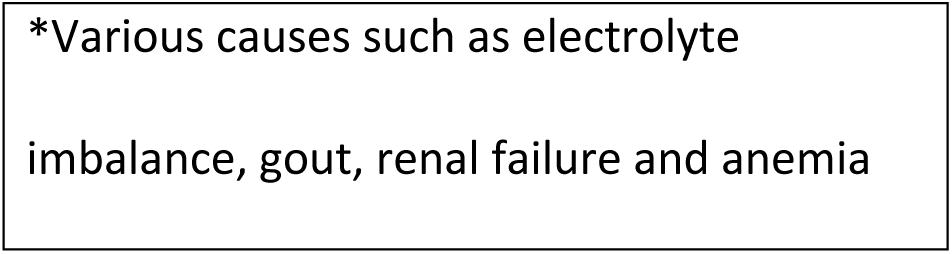
The most common causes for hospitalization during RM.

| Cause for hospitalization | (%) of all |
| --- | --- |
| Surgical procedure or trauma | 194 (29) |
| Infection | 77 (12) |
| Unspecified decline in<br>general condition | 61 (9) |
| Heart failure | 44 (7) |
| Myocardial ischemia | 43 (6) |
| Atrial arrhythmia | 42 (6) |
| COVID-19 | 29 (4) |
| Internal medicine* | 22 (3) |
| Syncope or presyncope | 21 (3) |
| Stroke/TIA | 16 (2) |
| **Total** | **549 (82)** |

\*Various causes such as electrolyte imbalance, gout, renal failure and anemia

Five hospitalizations (1%) were due to a device-related complication (three due to lead dislocation, one due to device infection and one due to pocket hematoma). Noteworthy, 21 % (145) of all hospitalizations were related to cardiovascular causes (heart failure, myocardial ischemia, stroke/TIA or arrythmias).

The number of ER evaluations, hospitalizations or the rate of ER evaluations or hospitalizations did not differ between the follow-up modalities. That is, 54 vs 50 patients had ER evaluation (p=0.6), 45 vs 36 patients were hospitalized (p=0.2). The rate of ER evaluations or hospitalizations did not differ between the follow-up methods, (IRR 1.2, 95% CI 0.8-1.7, p=0.3) and (IRR 1, 95% CI 0.8-1.4, p=0.8), respectively.

### Drop-out from RM

Since March 2024, only 3 patients have elected to discontinue RM. In one case the drop-out was due to advance dementia (hiding of the RM transmitter) and the other causes included afraid of electrical appliances and in the third case patient’s spouse complaint of hearing defect due to RM transmitter. The drop-out occurred six, eight and one month since the initiation of RM, respectively.

## Discussion

According to our real-word data of PM follow-up the RM-only approach reduced the workload of the pacemaker clinic without compromising patient safety. Due to the reduced need for in-office follow-up the RM-only approach burden patient less than appointment-based monitoring and interestingly - more than half of the patients followed by RM-only needed no in-office visits due to the pacemaker. The connectivity of RM was near-perfect, thus modern technology enables reliable RM-only approach.

In our center the follow-up of PM patients by RM-only started out as a natural experiment out of necessity. It rapidly gained positive feedback and over time it became a primary monitoring method in our center. Current expert statement on RM recommends scheduling an in-person visit every 24 months even for patients with continuous RM connectivity, and with no recent alerts or other cardiac comorbidity (15). However, if near-perfect connectivity, robust system to assure connectivity and excellent patient compliance exists the additional in-person visit may not be needed (15). In our cohort the success of daily transmissions was more than 90% and less than 10% (47/606) had success rate below 75%. It has not been defined what qualifies as near-perfect connectivity but in the appointment-based model, which remains the standard of care, the daily success rate for connectivity is just 0.3% when annual appointments are programmed. Our results indicate that sufficient connectivity can be achieved in a real-life setting with the current technology, which makes RM a compelling method for monitoring PM function. Unfortunately, our study focused only on Biotronik devices, so further studies are needed to confirm same level of connectivity in other systems.

Already in 2013, the results of the FOLLOWPACE study showed that many of the in-office PM follow-up visits were nonactionable, and it was suggested that most visits could be omitted or replaced by RM (^16^). It is tempting to speculate that in patients with efficient RM, also the in-office visits every 2 years would most likely be non-actionable. It should be highlighted that in-person visit burden patients. For example, a report from France showed that the median time due to a device clinic visit was 6 hours, and almost half of the patients needed a medical vehicle for travel ^(17)^. In the At-home study conducted in Japan the mean waiting time was one hour in the device clinic, the travelling time was more than thirty minutes, and most patients used car for transportation (^11^).

Thus, in-person visits are associated with cost to the patient and a carbon footprint for the environment. Noteworthy, time saved from travelling was considered the main cause for satisfaction with RM and majority of the patients were willing to replace the in-person visits with a remote follow-up (^17^). In our cohort more than half of the patients were followed solely by RM, meaning that no in-office visits due to the PM were needed. Nowadays, this proportion might be even higher (62%) as many of the reasons for the in-office visits in our study (*e.g*., interrogations due to MRI studies, lack of EGM in RM-portal after alert transmissions and forgetting to program RM on) have become obsolete with current technology. Although 25% of the device interrogations were found to be actionable, it is important to emphasize that many of the programming changes can be considered benign, for example, mode switches or adjustments to activity sensor settings. It is also unlikely that scheduled in-office appointments would have led to earlier detection of these issues, as most changes to activity sensor settings were made following patient requests and mode switch based on AF duration.

The safety of RM has been shown to be similar to the conventional monitoring combined with RM (^11, 12^), indicating that additional in-person visits do not improve patient safety. However, no randomized studies have directly compared exclusive RM versus the appointment-based care in PM patients.

Therefore, our results offer valuable new insights into the safety of exclusive RM and extend the findings of the AT-Home trial, which had a follow-up period limited to two years. It may be speculated that the RM-only approach could also be applied to patients with implantable cardioverter-defibrillators (ICDs), as many of these patients do not require pacing, making device function evaluation relatively straightforward.

Nevertheless, it must be kept in mind that this approach does not eliminate the need for regular follow-up of underlying cardiac and other comorbid conditions. In ICD patients, clinical follow-up could potentially be conducted in units without device interrogation capabilities—provided that an annual transmission is scheduled prior to the clinical appointment and the data is made available to the cardiologist responsible for the patient’s care.

Although the patients in this study were not randomized, the crossover design in a subgroup of patients minimizes confounding factors with the patients acting as their own control subjects. If anything, this method would bias against RM-only as during the RM-only period the patients were older and thus more likely to be hospitalized due various illnesses. Overall, it was seen that hospitalizations occur rarely in PM patients (0.3/year) and the causes are mainly related to other co-morbidities as specific cardiovascular etiology was present in one fifth of these patients.

The high volume of transmissions has been advocated as one of the disadvantages of RM and cited as one of the main reasons not to adopt RM (^18, 19^). It has been shown that patients randomized to RM are keener on requesting additional in-office follow-up than patient in standard care (^11, 12^). Thus, RM may increase the device clinic workload due to deluge of RM transmissions and subsequent additional in-person visits. However, we have previously shown that a hybrid strategy combining scheduled and alert transmission leads to a good balance of safety and RM workload (^14,20^). In a RM model based solely on alert transmissions it may be harder to justify turning alerts off for clinically less relevant events, leading to a higher burden of alerts and consequent increase in the workload. Compared to our data where an average of 1.3 RM transmissions were generated annually per device, in a contemporary report by O’Shea *et al*. the volume of mere alert transmissions in PM patients was 2 per device per year of which nearly 60% were due to high ventricular rate (HVR) or non-sustained ventricular tachycardia ^(18)^. In keeping with a previous study (^21^), less than half of the patients in our cohort had no alert transmissions during the follow-up. In our clinic scheduled transmissions are primarily evaluated by nurses experienced in RM, with infrequent need for electrophysiologist consultations. Therefore, it is likely that a nurse-driven hybrid approach may improve the safety without causing marked increase in the workload.

In our practice all patients receive a short and easily understandable summary of the alert and scheduled RM transmissions via a nationwide EHR system. This ensures the patients that their CIED has been evaluated and will likely improve patient satisfaction.

### Limitations

This study presents how follow-up by RM-only performs in a large real-world cohort of PM patients. However, certain limitations need to be addressed. The main limitation of this study is the non-random retrospective design. The RAPTOR-CIED study (NCT06937658) will offer future insights into alert-based remote monitoring, but currently, our study remains the only focused in this area.

Due to the design, it may be possible that patients with severe problems with RM may have dropped out before data collection and this may impact to the evaluation of RM performance between the year of initiation. However, the annual discontinuation rate in our cohort was extremely low.

The low dropout rate aligns with findings from earlier RM studies, where no crossover from the RM group to conventional care was observed (22). Interestingly, in our cohort, the dropouts were related to causes other than apparent safety concerns with RM.

The decision to include only patients with a Biotronik PM in the study is also a clear limitation. The rationale for this decision is described in the methods section. As the RM framework is quite similar with all pacemaker manufacturers, we believe that the main findings of this study e.g. the causes and occurrence for in-office visits during exclusive RM can be extrapolated to other manufacturers. Noteworthy, we are not aware of any studies, especially comparing the RM performance of different manufacturers and the differences between the vendors may complicate such studies. For example, only Biotronik allows the evaluation of daily success rate of transmissions and therefore in other vendors it is impossible to determine e.g. the duration of the possible disconnections. Additionally, the direct comparison of the occurrence of disconnection episodes is difficult as threshold is 14 days and unmodifiable in Medtronic and Boston and modifiable in Biotronik from 3 days to 21 days and Abbott from 7 days to 21 days. In our everyday work we have not noticed any meaningful differences between the vendors, but further studies are needed to verify, especially the performance of RM in other manufacturers.

It is possible that we did not catch all device interrogations, and thus underestimated the workload in RM. For example, interrogations in other hospital districts while the patient was travelling are not documented in our electronic patient records. However, an important feature of the Home Monitoring® is its ability to specify whether an in-office device interrogation has taken place, make such a bias unlikely. The workload of RM transmissions was not collected from the entire follow-up period and the volume of transmissions might have been higher in the beginning of the follow-up. The data of RM transmissions has been collected prospectively since 2023, and we believe this data is accurate and that the contemporary data is clinically more valuable as we have developed more pragmatic RM by analyzing this data (^14^).

Due to the retrospective nature of the study, we were unable to evaluate the mortality during RM as the patient cohort consisted only of patients in whom RM was active during data collection and the preservation of the data of deceased patients in Home-Monitoring® portal has been imperfect. However, in a previous study we showed that the deaths in a large cohort of RM patients were due to natural causes rather than related to RM (^14^). Thus, the mortality in a patient cohort with many other competitive causes of death may not necessarily be a reasonable endpoint to evaluate.

Finally, the emergence of conduction system pacing (CSP) may question the usefulness of RM-only approach, as verifying conduction system capture is difficult when relying only on real-time EGMs from RM portals. However, the loss on CSP during follow-up in relatively rare (^23^) and could be detected by combining the ECG evaluation with annual transmissions which can be evaluated through electronic health records.

## Conclusion

Our real-word data indicate that RM-only approach offers a feasible and safe method for long-term follow-up of pacemaker patients. As the number of PM implantations continues to rise, the role of the RM becomes increasingly important. RM-only approach reduced the workload compared to convention in-office follow-up of pacemaker patients without compromising safety. Hence, we believe that it may be time for a paradigm shift from appointment-based monitoring to remote-only monitoring in pacemaker patients.

## Clinical implications - Competency in Systems-Based Practice

Remote monitoring improves the safety of pacemaker follow-up. Our results indicate that remote monitoring can also be used as a primary follow-up method with clinic appointments programmed only when needed based on findings in remote monitoring.

This option could be offered to pacemaker patients which would change the current paradigm in pacemaker patient follow-up.

## Translational Outlook

Our results reinforce the benefits of remote monitoring in pacemaker patients follow-up but it is important to continue the development of remote monitoring to ensure that alert-based remote monitoring fulfills all the expectations of pacemaker patients concerning their follow-up.

## Data availability statement

The data that support the findings of this study are available from the corresponding author upon reasonable request.

## Abbreviations

CIED: Cardiac implantable electronic devices
RM: Remote monitoring
RM-only: Remote-only
PM: Pacemaker
HER: Electronic health record
HVR: High ventricular rate
AF: Atrial fibrillation
AHRE: Atrial high rate episode
ER: Emergency room
ICD: Implantable cardioverter-defibrillator

---

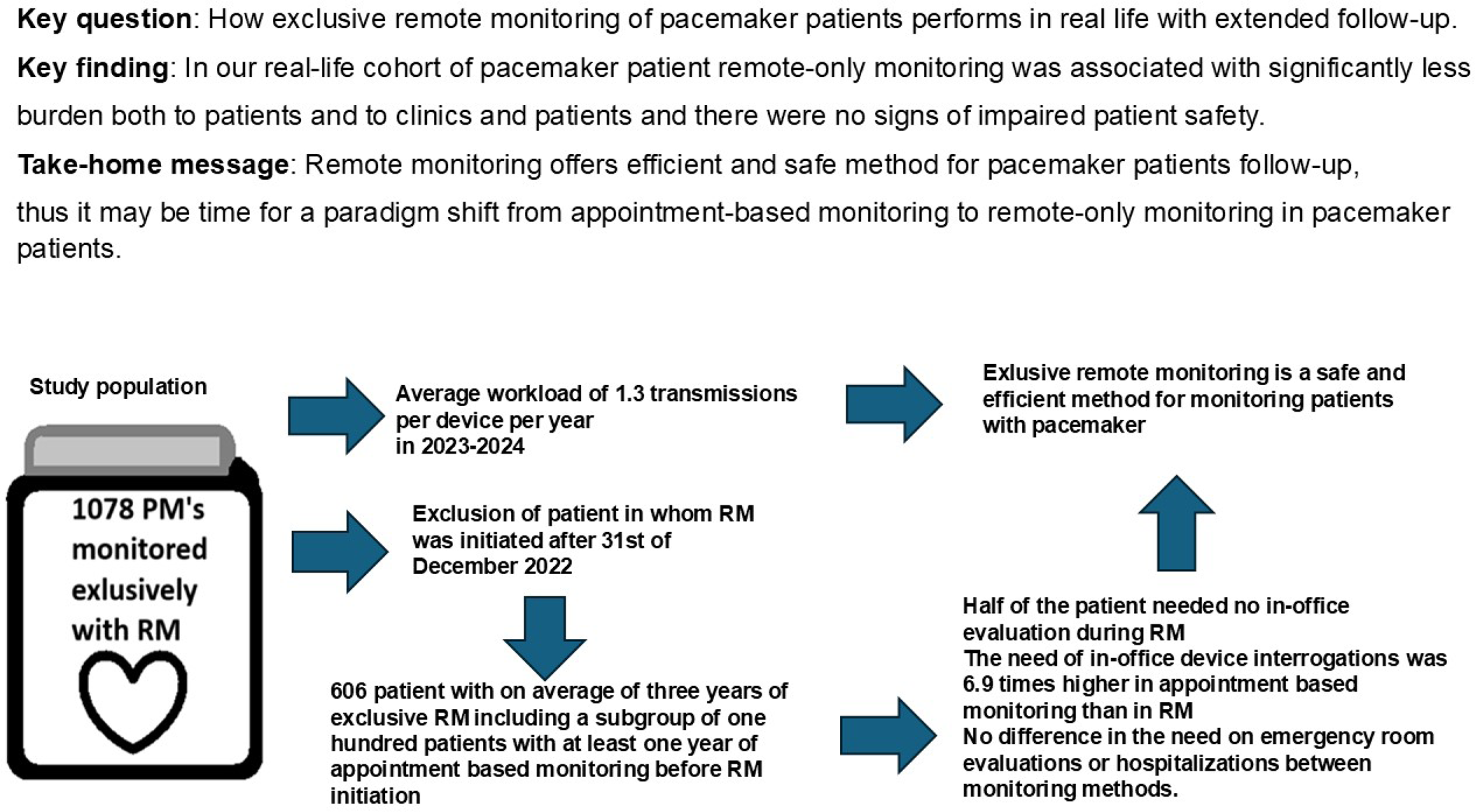

